# Starting at the community: Treatment seeking pathways of children with suspected severe malaria in Uganda

**DOI:** 10.1101/2021.12.09.21267055

**Authors:** Nina C. Brunner, Aliya Karim, Proscovia Athieno, Joseph Kimera, Gloria Tumukunde, Irene Angiro, Aita Signorell, Giulia Delvento, Tristan T. Lee, Mark Lambiris, Alex Ogwal, Juliet Nakiganda, Flavia Mpanga, Fred Kagwire, Maureen Amutuhaire, Christian Burri, Christian Lengeler, Phyllis Awor, Manuel W. Hetzel

**Author notes:** Corresponding authors: Nina Brunner,; Manuel Hetzel. Equal contribution (Phyllis Awor, Manuel W. Hetzel).

## Abstract

**Introduction:** Community health workers (CHW) usually refer children with suspected severe malaria to the nearest public health facility or a designated public referral health facility (RHF). Caregivers do not always follow this recommendation. This study aimed at identifying post-referral treatment-seeking pathways that lead to appropriate antimalarial treatment for children less than five years with suspected severe malaria.

**Methods:** An observational study in Uganda enrolled children below five years presenting to CHWs with signs of severe malaria. Children were followed up 28 days after enrolment to assess their condition and treatment-seeking history, including referral advice and provision of antimalarial treatment from visited providers.

**Results:** Of 2211 children included in the analysis, 96% visited a second provider after attending a CHW. The majority of CHWs recommended caregivers to take their child to a designated RHF (65%); however, only 59% followed this recommendation. Many children were brought to a private clinic (33%), even though CHWs rarely recommended this type of provider (3%). Children who were brought to a private clinic were more likely to receive an injection than children brought to a RHF (78% vs 51%, p<0.001) and more likely to receive the second or third-line injectable antimalarial (artemether: 22% vs. 2%, p<0.001, quinine: 12% vs. 3%, p<0.001). Children who only went to non-RHF providers were less likely to receive an artemisinin-based combination therapy (ACT) than children who attended a RHF (odds ratio [OR] = 0.64, 95% CI 0.51–0.79, p<0.001). Children who did not go to any provider after seeing a CHW were the least likely to receive an ACT (OR = 0.21, 95% CI 0.14–0.34, p<0.001).

**Conclusions:** Health policies should recognise local treatment-seeking practices and ensure adequate quality of care at the various public and private sector providers where caregivers of children with suspected severe malaria actually seek care.

## Introduction

Despite major reductions since the year 2000, malaria is still a leading cause of morbidity and mortality in children under 5 years of age, especially in Sub-Saharan Africa.^1^ Particularly children in remote areas with little access to health care are at increased risk of dying.^2 3^ To address this problem, the World Health Organization (WHO), the United Nations Children’s Fund (UNICEF) and others have developed an equity-focused strategy called integrated community case management (iCCM).^4^ Within the framework of iCCM programmes, community health workers (CHW) are trained to treat children less than 5 years with uncomplicated malaria, pneumonia and diarrhoea and to identify and refer severely ill children including those with fever and danger signs indicative of severe malaria. If available, CHWs may treat children with suspected severe malaria with a single dose of pre-referral rectal artesunate (RAS).^5^ The WHO treatment guidelines for malaria recommend that children who receive pre-referral treatment be referred to a health facility where appropriate post-referral treatment for severe malaria is available.^6^ The administration of the recommended post-referral treatment with an injectable antimalarial for at least 24 hours followed by a three-day course with an artemisinin-based combination therapy (ACT) depends on the confirmation of the severe malaria diagnosis by the post-referral provider. If confirmed, the WHO recommended parenteral antimalarial consists of intravenous or intramuscular artesunate. If artesunate is not available, severe malaria patients may be treated with intramuscular artemether or, as the least preferred option, intramuscular or intravenous quinine. However, the post-referral provider who re-assesses the child upon arrival at the referral facility may not always conclude that the child has severe malaria. If the post-referral provider diagnoses uncomplicated malaria, a three-day course with an ACT may be sufficient.

Locally adapted recommendations on appropriate referral facilities should ensure that children with suspected severe malaria are referred to a higher-level provider where appropriate treatment for severe malaria is available.^5^ CHWs are usually trained to refer to a health facility affiliated with the public health system, either the nearest or a designated referral facility.^7-14^ Previous studies found that CHWs normally referred in line with local referral guidance;^11 12^ yet, caregivers did not always follow the CHW’s referral recommendation. Factors that were negatively associated with referral completion were the administration of a pre-referral treatment, referrals on the weekend or at night, costs associated with seeking treatment, the distance to the recommended health facility, and perceived low quality of care at the recommended facility.^9 11 15-17^ On the other hand, children with signs of severe illness or perceived severity were more likely to complete referral.^9-12 16^ Alternative post-referral treatment seeking pathways often involved private providers.^11 12^ Whether non-adherence to official referral guidance and/or CHW’s referral advice led to inappropriate post-referral treatment was not part of these investigations. Therefore, these studies did not associate referral completion with an effectiveness measure of post-referral treatment seeking and treatment. To understand which post-referral treatment seeking pathways lead to appropriate and effective malaria treatment for children with suspected severe malaria, investigations of post-referral processes do not only need to take into consideration treatment seeking but the entire continuum of referral advise, adherence to advise, treatment seeking, and administration of post-referral treatment. A comprehensive descriptive approach allows recognizing shortfalls in treatment effectiveness along often complex treatment seeking pathways followed by children with suspected severe malaria.

In this study, we aimed at identifying treatment seeking pathways that led to the appropriate management of suspected severe malaria in children after initial treatment seeking from a CHW. We compared referral recommendations with actual post-referral treatment seeking actions to understand to what extent CHWs and caregivers deviated from the official recommendations. Finally, we described the reasons for selecting different types of post-referral providers to gain an insight into the motivations of caregivers to follow or disregard referral recommendations.

## Methods

### Study design

The data for this observational study was obtained from the Community Access to Rectal Artesunate for Malaria (CARAMAL) project, an operational research project accompanying the large-scale roll-out of RAS in Uganda, the Democratic Republic of the Congo, and Nigeria. In this analysis, we included only data from Uganda on disease episodes collected after the implementation of RAS during the period between April 2019 and August 2020. More details about the CARAMAL project and key results are described elsewhere (Lengeler et al. manuscript in preparation).^8 18 19^

### Study setting

The study area covered the districts of Kole, Kwania, and Oyam in the Lango region in the north of Uganda. In 2018, the total population of the three districts was almost 1 million of which around 20% were children under 5 years. In every village, two CHWs, locally called Village Health Team, provided close-to-home health care for children under 5 years with diarrhoea, pneumonia or malaria. The public health sector in the study districts consisted of 5100 CHWs (usually two per village), 30 primary health centres (PHC, locally called Health Centre II), and 20 referral health facilities (RHF) which provided free malaria diagnosis and treatment for children under the age of 5 years. CHWs and PHCs were community-based providers that were trained in the administration of pre-referral RAS, and instructed to refer the severe cases immediately to a designated RHF thereafter. RHFs were Health Centres III and IV or hospitals that had an inpatient ward and provided appropriate treatment for severe malaria including injectable antimalarials and ACTs. CHWs and public facilities were linked by a counter-referral system using a paper referral form to be taken to the post-referral provider by the caregiver and sent back to the CHW with information on final diagnosis and treatment.

### Data collection

CHWs enrolled children under 5 years with fever and at least one danger sign for which RAS was indicated as per Ugandan iCCM guidelines: unusually sleepy or unconscious, convulsions, inability to drink or feed anything, and persistent vomiting. For study purposes, CHWs tested eligible children with a malaria rapid diagnostic test (mRDT) since malaria testing of children with danger signs is not part of the iCCM algorithm. CHWs notified the local study team of the new enrolment via a text message service hosted by the Ugandan Ministry of Health (mTrac). CARAMAL study nurses called the CHWs to confirm the eligibility of the child and schedule a follow up visit at the child’s home 28 days after enrolment. In the case of death of the child the interview was scheduled two months later in observance of a mourning period. Study nurses were also present at three Health Centre IVs and one hospital in the study districts to record post-referral case management of community-enrolled children admitted to the inpatient ward. During follow up on day 28, caregivers were interviewed about signs and symptoms, treatment seeking history, diagnosis and treatment for their child’s illness episode. On the same occasion, information on the child’s condition and treatment administered was extracted from the CHW register.

CARAMAL study nurses collected data electronically using ODK Collect (https://opendatakit.org/) installed on Android tablets. At RHFs, data was first collected on paper before being entered into ODK Collect. The password protected ODK Aggregate server was hosted at the Swiss Tropical and Public Health Institute in Switzerland.

### Analysis

In this analysis, we included children who fulfilled the iCCM eligibility criteria for RAS administration, who were tested positive for malaria by any provider seen during the treatment seeking process, and for whom written informed consent was obtained. For children who passed, they were included if the death happened within 31 days after enrolment (28 days + 3 days in case a follow-up visit was delayed for operational reasons). Children who were brought to another provider before seeing a CHW, whose caregiver did not list the enrolling CHW during the follow up interview, and for whom more than 50% of the data on treatment was missing were excluded.

Treatment seeking pathways and the administration of antimalarials along the treatment seeking pathway were assessed based on the follow-up interviews with caregivers. Reliability of caregiver reported data was found sufficient when comparing to data obtained from provider records (Supplement Table S 1 and Table S 2). Health care providers were classified into RHFs, PHCs, CHWs, private clinics and drug shops. Antimalarial treatments of interest were RAS, antimalarial injections, and ACTs. For antimalarial injections, we used the administration of any injection as a proxy, after finding that receiving an injection according to the caregiver was predictive of receiving an injectable antimalarial according to RHF records (Supplement Table S 2). The same assumption was made for non-RHF providers after finding that PHCs, private clinics and drug shops rarely administered injections other than antimalarials (Supplement Table S 3). Antimalarial treatment was considered complete if a child received an ACT following an injectable treatment.

In addition, we assessed the health status of children at follow up (dead/alive), referral recommendations by CHWs and adherence to these recommendations, and reasons for seeking post-referral treatment. The latter information was only available if the interviewed person was the same person who brought the child to the post-referral provider.

For the treatment seeking pathways and malaria case management, we used the effectiveness decay model for community health interventions and its pathways representation as Sankey diagrams.^20^ To build the Sankey diagram, we used Tableau Public (https://public.tableau.com). Indicators were calculated as absolute numbers and proportions. Proportions were compared by chi-square test accounting for clustering at parish level. We used a random effects logistic regression model to investigate the association of post-referral treatment seeking with receiving an ACT and the health outcome of children. The models allowed for random intercepts for the parish of the home location of the child. Statistical analyses were conducted in Stata SE 16.1.

### Ethics

The CHWs informed caregivers about the CARAMAL study prior to enrolment and caregivers gave oral pre-consent to be contacted for a follow-up interview. We obtained written consent from all caregivers of provisionally enrolled children either at the RHF or before the follow-up interview 28 days after enrolment. This two-step consent procedure was selected to avoid interfering with the urgent referral process considering the severe nature of the child’s illness. The CARAMAL study protocol was approved by the Research Ethics Review Committee of the World Health Organization (WHO ERC, No. ERC.0003008), the Higher Degrees, Research and Ethics Committee of the Makerere University School of Public Health (No. 548), the Uganda National Council for Science and Technology (UNCST, No. SS 4534), and the Scientific and Ethical Review Committee of CHAI (No. 112, 21 Nov 2017). The study is registered on ClinicalTrials.gov (NCT03568344).

## Results

### Study population

Of the 2675 provisionally enrolled children, 2285 (85%) were successfully followed up and informed consent was obtained and 2245 (84%) met all inclusion criteria (Supplement Figure S 1). We excluded 34 children because of missing information or because they had been to another provider before going to the initial CHW. In total, 2211 children (83%) were included in this analysis.

The mean age of included children was 1.7 years (standard deviation [SD] = 1.2) and 1169 children (53%) were male (Table 1). The most common danger sign was being unusually sleepy or unconscious (88%); 42% of the children had convulsions. Interviewed caregivers were on average 29 years old (SD = 9.4) and the majority of them were female (N=1871, 85%). Inclusions were equally distributed across study districts.

**Table 1:** Characteristics of participating children and interviewed caregivers 28 days post-enrolment.

|  | N | % |
| --- | --- | --- |
| **Total** | **2211** |  |
| Child's mean age in years (SD) | 1.7 (1.2) |  |
| Child's sex |  |  |
| Male | 1169 | 52.9 |
| Female | 1042 | 47.1 |
| Danger signs |  |  |
| Convulsions | 927 | 41.9 |
| Unusually sleepy or unconscious | 1940 | 87.7 |
| Not able to drink or feed | 1696 | 76.7 |
| Vomiting everything | 1418 | 64.1 |
| Caregiver's mean age in years (SD) | 29.1 (9.4) |  |
| Caregiver's sex |  |  |
| Male | 340 | 15.4 |
| Female | 1871 | 84.6 |
| District |  |  |
| Kole | 753 | 34.1 |
| Kwania | 730 | 33.0 |
| Oyam | 728 | 32.9 |

### Sequence of providers

After seeing a CHW, almost all children (96%, N=2211) went to a second provider which was usually a public RHF (54%, N=2119) or a private clinic (33%, N=2119) (Table 2). Few children went to more than two providers (10%, N=2211). Most of them were children who had previously been to a RHF (69%, N=223) or to a PHC (23%, N=223). Of those who continued to at least a third provider and visited up to six providers, 126 (57%) went to a private clinic and 89 (40%) went to a RHF at some stage of the continued treatment seeking pathway.

**Table 2:** Treatment seeking and antimalarial treatment of children first attending a community health worker.

|  | First provider (CHW) |  | Second provider |  | Provider #3-6* |  |
| --- | --- | --- | --- | --- | --- | --- |
|  | N | % | N | % | N | % |
| **Total** | **2211** | **100** | **2119** | **95.8** | **223** | **10.1** |
| Treatment seeking |  |  |  |  |  |  |
| CHW | 2211 | 100 | 4 | 0.2 | 4 | 1.8 |
| PHC | NA |  | 189 | 8.9 | 2 | 0.9 |
| RHF | NA |  | 1149 | 54.2 | 89 | 39.9 |
| Private clinic | NA |  | 698 | 32.9 | 126 | 56.5 |
| Drug shop | NA |  | 79 | 3.7 | 7 | 3.1 |
| Case management |  |  |  |  |  |  |
| Received RAS | 1536 | 69.5 | 68 | 3.2 | 3 | 1.3 |
| Missing | 0 | 0.0 | 7 | 0.3 | 0 | 0.0 |
| Received injection | NA |  | 1168 | 55.1 | 132 | 59.2 |
| Missing | NA |  | 100 | 4.7 | 38 | 17.0 |
| Received ACT | 520 | 23.5 | 1426 | 67.3 | 100 | 44.8 |
| Missing | 22 | 1.0 | 102 | 4.8 | 39 | 17.5 |

\* Percentages add up to more than 100% because some children may have seen more than one provider
ACT= artemisinin-based combination therapy, CHW=community health worker, NA = not applicable, PHC = primary health centre, RAS=rectal artesunate, RHF= referral health facility

### Administration of antimalarial treatment

Among all children, 70% received RAS and 24% received an ACT from the enrolling CHW (Table 2). From the second provider, 1168 (55%) children received an antimalarial injection and 1426 (67%) received an ACT. Of the children receiving an injection, 534 (46%) received it from a RHF and 507 (43%) from a private clinic; but PHCs and drug shops also administered injections. AS illustrated in Figure 1, a variety of combinations of subsequent providers can result in a patient being treated according to guidelines with an injectable antimalarial and an ACT.

**Figure 1:**
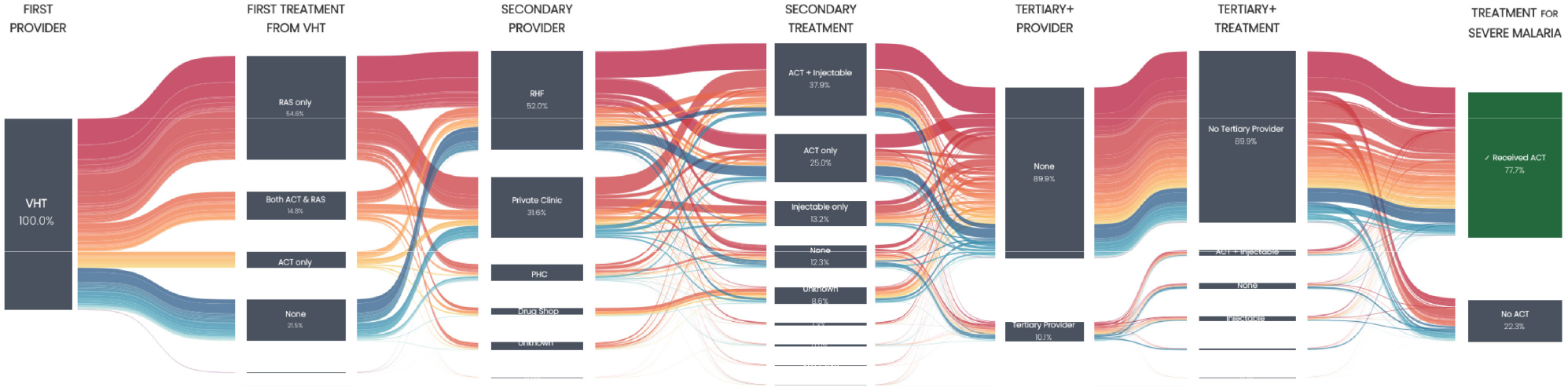
Sankey diagram of treatment seeking pathways and antimalarial treatment of children with suspected severe malaria first attending a community health worker. For technical reasons, some numbers may slightly differ from the numbers in the table and text.

Children who attended a private clinic as a second provider were more likely to receive an injection than children going to a RHF (78% vs 51%, p < 0.001), and among those receiving an injection, more likely to receive intramuscular artemether (22% vs. 2%, p<0.001) or injectable quinine (12% vs. 3%, p<0.001) (Supplement Table S 3). The majority of children that received an injection also received an ACT (866/1168=74%). Administration of an ACT without prior administration of an injectable was common among children going to a RHF (366/1149=32%) and those going to a PHC (74/189=39%). Meanwhile, 272 children (13%) did not receive any antimalarial treatment from the second provider (but they may have received different medication). These children more often continued to a third provider than children who had received antimalarial treatment at the second provider (47% vs. 5%). At the third and subsequent providers, both the administration of an injection (59%, N=223) and the administration of an ACT (45%, N=223) were common.

Taking into account the complete treatment seeking pathways documented in Figure 1, the administration of all three types of antimalarial treatments, RAS, injection and ACT, was common both for children who went to a RHF at some stage (41%, N=1192) and for children who never went to a RHF (40%, N=927) (Table 3). However, children that were brought to a RHF were more likely to receive an ACT than children seeking post-referral care from non-RHF providers (odds ratio [OR] = 0.64, 95% CI 0.51–0.79, p<0.001). Children who did not go to any provider after seeing a CHW were the least likely to receive an ACT (OR = 0.21, 95% CI 0.14–0.34, p<0.001). In addition, they were more likely to die than children going to a RHF (OR = 5.97, 95% CI 1.80–19.78, p=0.003). Children who went to a non-RHF provider were less likely to die than children who had been to a RHF, but the difference was not statistically significant (OR = 0.14, 95% CI 0.02–1.12, p=0.06).

**Table 3:** Antimalarial drug administration if children went to referral health facility (RHF) at any stage of the treatment seeking pathway, went to at least one other provider which was not a referral health facility (non-RHF provider), or did not go to any other provider after seeing the first provider.

|  | RHF |  | Non-RHF provider§ |  | No provider |  | Total |  |
| --- | --- | --- | --- | --- | --- | --- | --- | --- |
|  | N | % | N | % | N | % | N | % |
| All | 1192 | 100 | 927 | 100 | 92 | 100 | 2211 | 100 |
| Case management* |  |  |  |  |  |  |  |  |
| No antimalarial treatment | 62 | 5.2 | 43 | 4.6 | 7 | 7.6 | 112 | 5.1 |
| RAS only | 64 | 5.4 | 46 | 5.0 | 38 | 41.3 | 148 | 6.7 |
| Injection only | 23 | 1.9 | 28 | 3.0 | 0 | 0.0 | 51 | 2.3 |
| ACT only | 180 | 15.1 | 85 | 9.2 | 15 | 16.3 | 280 | 12.7 |
| RAS + injection | 64 | 5.4 | 118 | 12.7 | 0 | 0.0 | 182 | 8.2 |
| RAS + ACT | 212 | 17.8 | 129 | 13.9 | 32 | 34.8 | 373 | 16.9 |
| Injection + ACT | 104 | 8.7 | 104 | 11.2 | 0 | 0.0 | 208 | 9.4 |
| RAS + injection + ACT | 483 | 40.5 | 374 | 40.3 | 0 | 0.0 | 857 | 38.8 |
| Received ACT | 979 | 82.1 | 692 | 74.6 | 47 | 51.1 | 1718 | 77.7 |
| Dead at follow-up | 9 | 0.8 | 1 | 0.1 | 4 | 4.3 | 14 | 0.6 |
|  | **OR** | **95% CI** | **OR** | **95% CI** | **OR** | **95% CI** |  |  |
| Received ACT | Ref. |  | 0.64 | 0.51-0.79 | 0.21 | 0.14-0.34 |  |  |
| Dead at follow-up | Ref. |  | 0.14 | 0.02-1.12 | 5.97 | 1.80-19.78 |  |  |

\* By any provider along the treatment seeking pathway
§ CHWs, PHCs, private clinics, or drug shops
ACT= artemisinin-based combination therapy, CHW=community health worker, PHC = primary health centre, RAS=rectal artesunate, RHF= referral health facility

### Referral recommendations and adherence

For most children (93%, N=2211), caregivers received a referral recommendation from the CHW (Table 4). The majority of CHWs recommended the caregiver to take their child to a RHF (65%). We could not determine the exact type of recommended post-referral provider for 344 (16%) children because of a lack of detail in the data. Few children were sent to a PHC (9%), a private clinic (3%), or a drug shop (<1%). No CHW recommended referral to another CHW. Overall, adherence to the CHWs’ recommendations was 60% (N=1712). Adherence was comparable for children sent to a RHF (59%, N=1439) or to a PHC (59%, N=204), but was significantly higher among children sent to a private clinic (82%, N=65, p<0.001). The majority of children who did not adhere to the advice to go to a RHF or PHC went to a private clinic (463/593=78% and 47/84=46%, respectively) (Figure 2).

**Table 4:** Referral recommendation of the first CHW provider and referral adherence to referral recommendation.

|  | Referral recommendation |  | Adherence with recommendation |  |
| --- | --- | --- | --- | --- |
|  | N | % | N | % |
| **Total** | **2211** | **100** |  |  |
| None | 155 | 7.0 |  |  |
| Provider | **2056** | **93.0** | **1712** | **59.7\*\*** |
| CHW | 0 | 0.0 | NA |  |
| PHC | 204 | 9.2 | 120 | 58.8 |
| RHF | 1439 | 65.1 | 846 | 58.8 |
| Private clinic | 65 | 2.9 | 53 | 81.5 |
| Drug shop | 4 | 0.2 | 3 | 75.0 |
| Unknown facility type* | 344 | 15.6 | NA |  |

\* PHC, RHF or private clinic
\*\* N=1712, excluding children recommended to go to an unknown facility type
CHW=community health worker, NA = not applicable, PHC = primary health centre, RAS=rectal artesunate, RHF= referral health facility

**Figure 2:**
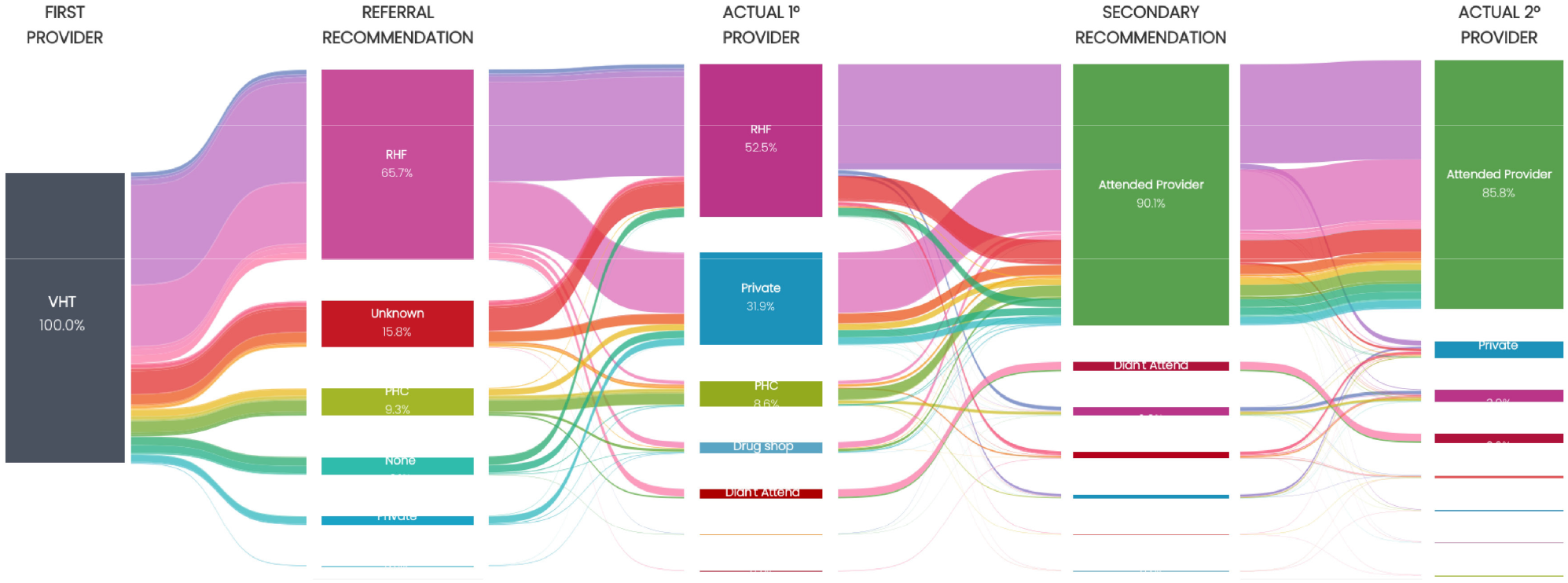
Sankey diagram of referral recommendations and actual treatment seeking of children with suspected severe malaria first attending a community health worker. For technical reasons, some numbers may slightly differ from the numbers in the table and text.

### Reasons for going to post-referral provider

Most children who were brought to a RHF as a second provider (N=1099) were taken there because the previous provider, i.e. the CHW, had referred them there (83%) (Table 5). The same was true for the 179 children going to a PHC (87%). Besides being referred, the experience and professionalism of the provider were frequently mentioned reasons to attend a RHF (29%) or a PHC (31%). In contrast, being referred was substantially less frequently mentioned for the 659 children brought to a private clinic (24%). Meanwhile, knowing the provider was an important reason to go to a private clinic (49%). As for PHCs and RHFs, experience and professionalism of the provider played an important role (35%). Reasons for going to a third or subsequent provider were largely the same, except for children more often going to a private provider because they were referred there (39%, N=126) (Supplement Table S 4).

**Table 5:** Reasons for going to second provider after attending a community health worker (CHW), by type of second provider.

|  | CHW |  | PHC |  | RHF |  | Private clinic |  | Drug shop |  |
| --- | --- | --- | --- | --- | --- | --- | --- | --- | --- | --- |
|  | N | % | N | % | N | % | N | % | N | % |
| **Total** | **4** | **100** | **179** | **100** | **1099** | **100** | **659** | **100** | **73** | **100** |
| Reason |  |  |  |  |  |  |  |  |  |  |
| Knowing the provider | 1 | 25.0 | 20 | 11.2 | 136 | 12.4 | 321 | 48.7 | 0 | 0.0 |
| Trusting the provider | 2 | 50.0 | 11 | 6.1 | 102 | 9.3 | 78 | 11.8 | 6 | 8.2 |
| Experience and professionalism | 1 | 25.0 | 55 | 30.7 | 318 | 28.9 | 230 | 34.9 | 41 | 56.2 |
| Custom | 0 | 0.0 | 17 | 9.5 | 103 | 9.4 | 25 | 3.8 | 0 | 0.0 |
| Low costs | 2 | 50.0 | 7 | 3.9 | 84 | 7.6 | 120 | 18.2 | 12 | 16.4 |
| Referred by other provider | 0 | 0.0 | 156 | 87.2 | 910 | 82.8 | 156 | 23.7 | 6 | 8.2 |

CHW=community health worker, PHC = primary health centre, RAS=rectal artesunate, RHF= referral health facility

## Discussion

Treatment guidelines for severe malaria emphasize the importance of appropriate post-referral treatment to children with suspected severe malaria after the administration of pre-referral treatment by a community-based provider. Currently, the scientific literature lacks comprehensive analyses that relate post-referral treatment seeking to the appropriateness and effectiveness of post-referral antimalarial treatment. In this study, we investigated treatment seeking decisions taken by caregivers of a child with suspected severe malaria who attended a CHW in Northern Uganda. We focused on antimalarial treatment administered along the treatment seeking pathway.

After initial treatment seeking from a CHW, almost all children went to a second provider. While the majority of CHWs recommended referral to a RHF, private clinics were a common choice for post-referral treatment. Private clinics were often visited as a second provider (32.9%) even though CHWs rarely recommended referral to a private clinic (2.9%). In case a CHW did recommend referral to a private clinic, more caregivers followed their recommendation compared the recommendation to go to either an RHF or PHC. In private clinics, children were more likely to receive an injection than in RHFs where quite often only an ACT was given. Overall, the proportion of children receiving an ACT was high (77.7%); however, children who never went to an RHF were significantly less likely to get an ACT. Most caregivers who brought their child to an RHF did so due to the CHW’s referral recommendation. Knowing the provider was the predominant reason for caregivers who brought their child to a private clinic.

According to the results of this study, general access to formal health care services did not appear to be a problem in the study area; but quality of care for severe malaria patients should be improved. The finding of certain children, particularly those never attending a formal RHF, being treated with an artemisinin monotherapy is concerning as it may contribute to the selection of artemisinin resistance parasites. The establishment of artemisinin-resistant *P. falciparum* in Northern Uganda has recently been documented and the selection of resistant strains was found to be associated with substandard treatment practices (Awor et al. manuscript in preparation).^21^ An urgent response is needed to mitigate the spread of resistance to the current first-line treatments for malaria. Based on the findings of this study, we suggest a number of complementary targeted interventions aiming at improving quality of care for children with suspected severe malaria while at the same time taking into consideration the local landscape of treatment seeking pathways.

First, increased efforts to assure completion of antimalarial treatment with an ACT are particularly important in the private sector. In the present study, children who never went to a RHF and did not receive an ACT were mainly children attending private clinics. However, the role of the private sector as a source of post-referral treatment is paradoxical. On the one hand, referral to a private clinic may not be recommendable because of concerns around the quality of care,^22^ mainly the non-conformity with guidelines, lack of training and the availability of poor-quality drugs.^23-28^ The advantage of training CHWs to refer to a health facility affiliated with the public health system is that official health authorities can control the level of training of the higher-level provider, and, in the case of a referral facility, the availability of appropriate and quality-assured severe malaria treatment. Additionally, higher-level facilities often provide supervision and drugs to CHWs,^4 29^ meaning that there is an established link between both types of providers, which may facilitate referral processes.^30^ On the other hand, private facilities are often more accessible than public facilities,^22 31 32^ and their presence can improve access to health care for children who would otherwise be left untreated.^8^ Additionally, the results of this study suggest that RHFs in some instances referred children to private clinics where at least some of them must have received an ACT. The reason for further referral may have been ACT stock-outs at the RHF.^33 34^ Consequently, the private sector would in fact contribute to appropriate post-referral treatment for children who first complete referral to a RHF. However, there was also anecdotal evidence of health workers at RHFs sometimes referring patients to their private drug shop or clinic, despite available drug stocks at the RHF. Similar reasons for referral have previously been reported in Tanzania.^33^ In the fight against artemisinin resistance, strategies to support health workers’ compliance with guidelines and policies are crucial in both the public and private sector.

Second, follow-ups of children by CHWs only hours after referral should be encouraged in order to ensure completion of malaria treatment with an ACT.^5^ There were few children that did not go to any provider other than the initial CHW, but their chance of not receiving an ACT was substantially higher compared to children attending a second health care provider. A timely follow-up of these children at home would allow the CHW to confirm if the referral recommendation was followed and take appropriate action. The latter could include reinforcing the referral recommendation, the provision of a second dose of RAS (as per current WHO recommendation in case of referral delay), or the provision of an oral ACT if referral was considered impossible and the child’s condition had already improved sufficiently. Importantly, sufficient stocks of RAS and ACTs are a prerequisite for a functioning follow-up system that may contribute to increasing the proportion of children receiving a full course of ACT in spite of failing to seek post-referral treatment from a RHF. Children who did not seek post-referral treatment were also more likely to die. However, this finding should be interpreted with caution as it may include children who died shortly after being attended by a CHW with no time to seek further care.

Third, the counter-referral system, where referral forms are returned to the CHW should be strengthened. In addition to ensuring post-treatment data collection, information on the final diagnosis and treatment provided by RHFs may provide an improved learning experience for CHWs, which may eventually lead to an improvement in distinguishing non-severe from severe cases. In this study, several children were treated only with an ACT (no injectable antimalarial) at the post-referral provider, especially at RHFs, suggesting that these children could already tolerate oral antimalarial treatment. While the completion of referral is critical if the child requires injectable antimalarial treatment or professional management of complications, there may be a window of opportunity for treating certain children with an oral antimalarial after a first dose of RAS even though they were initially found with danger signs of severe illness. Such treatment may not necessarily have to be provided at a RHF. Reducing referrals of less severely ill children can contribute to decreasing out-of-pocket expenditures related to health care seeking.^34^

It is important to note that follow-ups of children at home to verify referral and to complete the counter referral process lead to a higher workload for CHWs. Even though CHWs are often community members without formal work engagements, taking care of the community’s sick children comes with opportunity costs for taking care of their own households (child care, farming etc.).^35 36^ To compensate CHWs for their investments, recommendations to formalize iCCM programmes and pay CHWs have been made in the past and emphasized again more recently.^37 38^ In Uganda, attempts into this direction have been made; however, so far with little success.^39^ Considering the deprived state of remote communities, strategies to improve access should not be to the disadvantage of those they are claiming to help.

Fourth, investments into the supply side should be complemented by the removal of barriers to appropriate treatment on the demand side. Effective strategies may be educating caregivers on the importance of ACT administration after injectable antimalarials,^40^ and making antimalarials more affordable in the private sector. In Uganda, ACTs are subsidized through the private sector co-payment mechanism (CPM).^27^ However, the funding for CPM has been reduced significantly since 2017.^41^ In 2018, quality-assured ACTs in the private sector were more expensive than non-quality-assured ACTs and non-subsidized injectable artemether.^27 41^ Considering that caregivers in resource-limited settings are price-sensitive,^27^ the comparatively high price for ACTs may explain why injectable antimalarials were not followed by the administration of an ACT, provided that ACTs were available and health workers in the private sector were sufficiently trained.

Furthermore, it may be worthwhile considering the regulation of intramuscular antimalarials at PHC level and in the private sector. In Uganda, the use of injectable antimalarials is currently not recommended below Health Centre III level.^42^ However, findings of this study and anecdotal evidence suggest that the administration of intramuscular and potentially intravenous antimalarials was a common practice in PHCs and in private clinics. The use of intramuscular artesunate or artemether has previously been proposed in Ethiopia and Nigeria with ambiguous conclusions on the feasibility of the approach.^43 44^ Both studies suggested that implementation research could evaluate whether such an approach is feasible and effective in treating children with moderately severe malaria at lower-level facilities.

The findings of this study are limited by the reliability and completeness of caregiver-reported data. The proportion of children receiving an ACT from a CHW or RHF as reported by the caregivers exceeded the proportions reported by these providers. Apart from caregiver recall bias, it is possible that some treatments were not recorded by CHWs or at RHFs. Agreement between caregiver reported ACT administration and ACT administration recorded at the RHF was higher if prescribed treatments were taken into account (Supplement Table S 2). This finding suggests that ACT prescriptions were usually filled in. However, the risk of overestimating ACT administration remains. Conversely, children with missing data on ACT administration were considered as not having received an ACT, which may have led to an underestimation of the proportion of children receiving an ACT.

## Conclusion

Achieving universal health coverage including access to effective and affordable treatment is one of the most important components of the Sustainable Development Goal 3. To attain this target, it is crucial to understand the actions taken and constraints faced by both patients and health care providers, as a basis for targeted improvements. New policies and recommendations should recognise existing systems and practices, as illustrated here comprehensively for one region in Uganda, so that targeted interventions are implemented where they are most needed and likely to have the highest impact.

## Data Availability

The data that support the findings of this study are available from the corresponding author upon reasonable request.

## Statements

## Acknowledgements

The authors would like to sincerely thank all caregivers and children who participated in this study, health care providers and local health authorities who facilitated data collection activities, all data collectors who generated the evidence for this study, and colleagues at Clinton Health Access Initiative (CHAI), Unicef, and the Uganda Ministry of Health, particularly Jimmy Opigo, for their support. We appreciate Nadja Cereghetti’s support to the management of the CARAMAL Project and thank Aurelio Di Pasquale for setting up and maintaining the ODK Aggregate server.

## Competing interests

Authors received financial support for this study from Unitaid. Authors declare no financial relationships with any organizations that might have an interest in the submitted work in the previous three years; no other relationships or activities that could appear to have influenced the submitted work.

## Funding

This study was funded by Unitaid.

## Notes

### Competing Interest Statement

The authors have declared no competing interest.

### Clinical Trial

NCT03568344

### Funding Statement

This study was funded by Unitaid (grant reference XM-DAC-30010-CHAIRAS).

### Author Declarations

The Research Ethics Review Committee of the World Health Organization (WHO ERC, No. ERC.0003008), the Higher Degrees, Research and Ethics Committee of the Makerere University School of Public Health (No. 548), the Uganda National Council for Science and Technology (UNCST, No. SS 4534), and the Scientific and Ethical Review Committee of CHAI (No. 112, 21 Nov 2017) gave ethical approval for this work.

