## Supplementary materials for "Starting at the community: Treatment seeking pathways of children with suspected severe malaria in Uganda"

*Table S 1: Positive and negative predictive values of caregiver reported administration of antimalarial treatment with administration according to the enrolling community health worker (CHW) as reference. A) Child received rectal artesunate (RAS) according to the caregiver versus received RAS according to the enrolling CHW. B) Received an artemisinin-based combination therapy (ACT) according to the caregiver versus received an ACT according to the enrolling CHW.*

A)

| | | **Enrolling CHW** | | | |
|---|---|---|---|---|---|
| | | Received RAS | Did not receive RAS | Predictive values | |
| **Caregiver** | Received RAS | 1495 | 29 | Positive: | 98% |
| | Did not receive RAS | 42 | 618 | Negative | 94% |

B)

| | | **Enrolling CHW** | | | |
|---|---|---|---|---|---|
| | | Received ACT | Did not receive ACT | Predictive values | |
| **Caregiver** | Received ACT | 400 | 114 | Positive: | 78% |
| | Did not receive ACT | 138 | 1511 | Negative | 92% |

*Table S 2: Positive and negative predictive values of caregiver reported administration of antimalarial treatment with administration according to referral health facility records as reference. A) Child received any injection according to the caregivers versus received an injectable antimalarial according to the facility records. B) Child received an artemisinin-based combination therapy (ACT) according to the caregiver versus received an ACT according to facility records. C) Child received an ACT according to the caregiver versus received or was prescribed an ACT according to facility records.*

A)

| | | **Referral health facility** | | | |
|---|---|---|---|---|---|
| | | Received an injectable antimalarial | Did not receive an injectable antimalarial | Predictive values | |
| **Caregiver** | Received an injection | 126 | 4 | Positive: | 97% |
| | Did not receive an injection | 3 | 1 | Negative | 25% |

B)

| | | **Referral health facility** | | | |
|---|---|---|---|---|---|
| | | Received ACT | Did not receive ACT | Predictive values | |
| **Caregiver** | Received ACT | 38 | 87 | Positive: | 30% |
| | Did not receive ACT | 4 | 5 | Negative | 56% |

C)

| | | **Referral health facility** | | | |
|---|---|---|---|---|---|
| | | Received or was prescribed ACT | Did not receive and was not prescribed ACT | Predictive values | |
| **Caregiver** | Received ACT | 119 | 1 | Positive: | 99% |
| | Did not receive ACT | 3 | 2 | Negative | 40% |

*Table S 3: Type of injections administered by the second provider, by type of second provider.*

| | PHC | | RHF | | Private clinic | | Drug shop | |
|---|---|---|---|---|---|---|---|---|
| | N | % | N | % | N | % | N | % |
| Received injection | 44 | 100 | 558 | 100 | 517 | 100 | 49 | 100 |
| Type of injectable drug* | | | | | | | | |
| Artesunate | 18 | 40.9 | 462 | 82.8 | 142 | 27.5 | 15 | 30.6 |
| Artemether | 2 | 4.5 | 9 | 1.6 | 111 | 21.5 | 20 | 40.8 |
| Quinine | 6 | 13.6 | 19 | 3.4 | 62 | 12.0 | 5 | 10.2 |
| Other | 4 | 9.1 | 40 | 7.2 | 18 | 3.5 | 1 | 2.0 |
| Don't know | 18 | 40.9 | 74 | 13.3 | 205 | 39.7 | 10 | 20.4 |

* Administration of multiple injectable drugs is possible

*Table S 4: Reasons for going to the third or any other provider thereafter, by type of provider.*

| | CHW | | PHC | | RHF | | Private clinic | | Drug shop | |
|---|---|---|---|---|---|---|---|---|---|---|
| | N | % | N | % | N | % | N | % | N | % |
| **Total** | **4** | **100** | **2** | **100** | **89** | **100** | **126** | **100** | **7** | **100** |
| Reason | | | | | | | | | | |
| Knowing the provider | 0 | 0.0 | 0 | 0.0 | 14 | 15.7 | 46 | 36.5 | 0 | 0.0 |
| Trusting the provider | 0 | 0.0 | 0 | 0.0 | 10 | 11.2 | 13 | 10.3 | 0 | 0.0 |
| Experience and professionalism | 1 | 25.0 | 0 | 0.0 | 35 | 39.3 | 49 | 38.9 | 5 | 71.4 |
| Custom | 0 | 0.0 | 1 | 50.0 | 13 | 14.6 | 10 | 7.9 | 1 | 14.3 |
| Low costs | 1 | 25.0 | 0 | 0.0 | 8 | 9.0 | 19 | 15.1 | 1 | 14.3 |
| Referred by other provider | 0 | 0.0 | 2 | 100.0 | 64 | 71.9 | 49 | 38.9 | 1 | 14.3 |

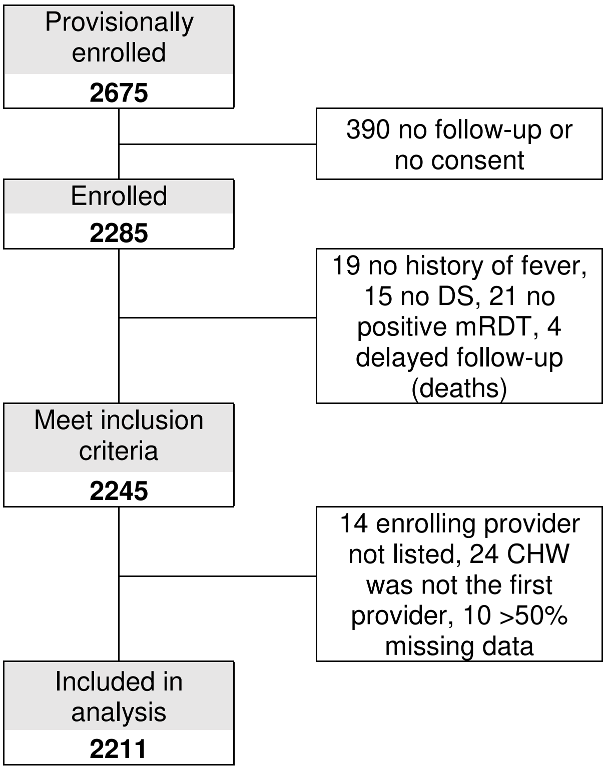


*Figure S 1: Flowchart*
